# Using supermarket loyalty card data to investigate seasonal variation in laxative purchases in the UK

**DOI:** 10.1101/2025.08.01.25332648

**Authors:** Romana Burgess, Neo Poon, Edward Sloan, James Goulding, Helen Bould, Anya Skatova

## Abstract

While laxatives are designed to manage the symptoms of constipation, they are also frequently misused for weight control, particularly by individuals with eating disorders. This study aims to investigate the relationship between laxative purchases and weight management by examining seasonal trends in laxative purchasing. Using real-world transaction data from a major UK pharmacy retailer spanning December 2013 to December 2014, we analyse purchasing patterns from 748,375 buyers to explore potential links with weight control behaviours.

In pre-registered analysis, we use regression models to investigate our hypotheses: that the number of doses purchased would be greater in January compared to the previous December (reflecting motivations in relation to “New Year’s resolutions” around weight loss), and greater in May-August compared to the subsequent September (reflecting an increased focus on body image during the summer). We examine differences between stimulant and non-stimulant laxatives, as stimulants are more commonly misused for weight control due to their rapid effects. To validate our findings, we compare purchasing patterns with those for diet and weight management products over the same periods, and also include negative controls of unrelated medication groups (i.e., painkillers, cold and flu, and hay fever medications).

Our findings reveal seasonal variations in laxative purchasing, particularly for non-stimulant medications. Purchases increase in January compared to December and are higher in some summer months compared to September, suggesting potential use for weight control. Non-stimulants exhibit greater seasonal fluctuation than stimulants. Purchase of weight management products followed similar patterns, aligning with established seasonal trends in weight loss behaviours. Laxative purchases show seasonal trends aligned with other weight management produce purchases, potentially aligned to use motivated by body image concerns. These findings provide retrospective support for the MHRA stimulant laxative policy change and highlight the need for further evaluation of its impact.

**Author Summary:** Laxatives are commonly used to relieve constipation, but they can also be misused by people trying to lose weight—particularly those struggling with eating disorders. We wanted to understand whether people in the UK might be using laxatives for weight control at certain times of the year. To do this, we analysed loyalty card data from a large UK pharmacy to look at when people bought laxatives over a one-year period.

We found that purchases of laxatives increased during the summer months, a time when many people focus on their appearance and may try to lose weight because of the warmer weather. We also saw similar patterns in purchases of diet-related products, which supports the idea that laxatives may sometimes be used for weight management.

Our research shows how loyalty card data can offer insights into health behaviours in the general population. These findings could help inform public health efforts and policy, especially around eating disorders and the regulation of over-the-counter medications. This approach could also be applied to other types of health-related behaviours, offering a new way to support early identification and intervention.

## 1. Introduction

Laxatives are medications used to manage constipation, readily available to buy over-the-counter in the UK. The prevalence of the use of such medication varies widely, ranging from 1% to 18% in the general population and from 3% to 59% among those experiencing constipation [1]. Laxatives however, are also commonly (mis)used by individuals with eating disorders as an unhealthy weight-control behaviour, with clinical studies reporting usage rates as high as one in four [2, 3]. The widespread misuse among individuals with eating disorders poses significant public health concerns [4, 5]. Particularly alarming is the misuse of stimulant laxatives, which are associated with severe health risks, including organ damage, electrolyte imbalances, and long-term gastrointestinal complications [6]. Stimulant laxatives are frequently misused, likely due to their fast-acting effects, which may reinforce the misguided belief that they can prevent calorie absorption by inducing diarrhoea [7].

Understanding laxative misuse amongst individuals with eating disorders—and more broadly, laxative use within the general population [1]—has primarily relied on self-reported data [2, 3], which is subject to recall errors and social desirability effects [8]. Given the stigma attached to such usage, loyalty card data promises a novel and relatively unexplored method for tracking self-medication, having only been applied in this setting in a handful of studies and never at national scale. Previous studies have explored the predictive power of over-the-counter medication sales in assessing common ailments in the UK: pain relief and cough and cold medicines have been directly linked to respiratory mortality rates at local levels [9]; the impact of demographics in predicting treatment of minor ailments has been examined, alongside sun preparation sales, with men making far fewer purchases than women [10]; and purchases of “symptomatic” over-the-counter medications (e.g., indigestion medicines) have been shown to be indicative of subsequent ovarian cancer diagnoses [11]. Despite offering an objective measure of self-medication, no studies have yet used loyalty card transactions to track laxative self-medication—a potential gap given the significant risks associated with misuse of such products. This study addresses that gap by exploring patterns in laxative self-medication, with a focus on variations during the New Year period and over the summer months in order to investigate instances where changes in use might be related to weight loss attempts.

### 1.1. Hypotheses

Loyalty card data are examined to identify seasonal patterns in laxative purchasing, hypothesising that stimulant laxative purchases in relation to a wish to lose weight will display two primary seasonal peaks: one following the festive season—reflecting motivations in relation to “New Year’s resolutions” around weight loss—and another during the summer—a period often associated with heightened attention to appearance due to the warmer weather.

For the New Year period, we propose that:

- **H1a)** the number of stimulant laxative doses purchased will be higher in January than in the December prior;
- **H2a)** this seasonal variation will be more evident for stimulant laxatives than for other types of laxatives;
- **H3a)** this seasonal variation will be also observed in “diet and weight management” products sold at the same retailer.

We anticipate a second seasonal increase in the warmer months, potentially related to an increased focus on appearance. We hypothesise that:

- **H1b)** the number of stimulant laxative doses purchased will be higher in May, June, July, and August than in September;
- **H2b)** this seasonal variation will be more evident for stimulant laxatives than for other types of laxatives;
- **H3b)** this seasonal variation can also be observed in “diet and weight management” products sold at the same retailer.

We expect that laxative use for constipation will show little seasonal variation, as constipation is well evidenced to not vary seasonally [12]. However, we do expect laxative use with the goal of weight loss to vary seasonally in line with other indicators of attempted weight loss (e.g., purchasing fewer calories) [13]. As such, we assume that seasonal variation in laxative purchases is likely related to their use for attempted weight loss.

Negative controls of other medication groups are used to observe whether seasonal variation is due to other unrelated factors. That is, we expect that other groups of medication—pain relief, hay fever and allergies, cold and flu—will not show the same seasonal pattern in sales, testing whether patterns are driven by external factors such as introduction of new products, or broader seasonal variation in purchasing over the counter medications. With pain relief, we do not expect any seasonality; with hay fever, we expect increases in spring or early summer due to common rise of pollen-induced allergy symptoms; and with cold and flu we expect an increase in colder months.

## 2. Results

### 2.1. Descriptive Summary

Data from the top 10% of laxative buyers were used within our analyses, representing 73,742 customers each purchasing at least 60 doses-worth of medication between December 2013 and December 2014. This accounted for 726,357 items purchased across 638,318 transactions. The mean number of doses in a single stimulant product was 21.31 (SD = 15.52) (i.e., a stimulant product would last maximum 21 days, if taking one dose per day), while the mean number of doses in a single non-stimulant product was 8.93 (SD = 6.11) (i.e., a non-stimulant product would last maximum 9 days on average).

#### 2.1.1. Monthly Product Sales

Mean product sales per month were calculated between December 2013 and December 2014 for the top 10% of buyers in five product categories—all laxatives (stimulant and non-stimulant) (n = 85,578 buyers), pain killers (n = 553,414), weight management (n = 109,827), hay fever (n = 236,085), and cold/flu medications (n = 469,651). This is shown in Figure 1. Note that actual sales units (i.e., number of products sold) were used here, as it was infeasible to manually translate all products across all categories into dosages.

**Fig 1:**
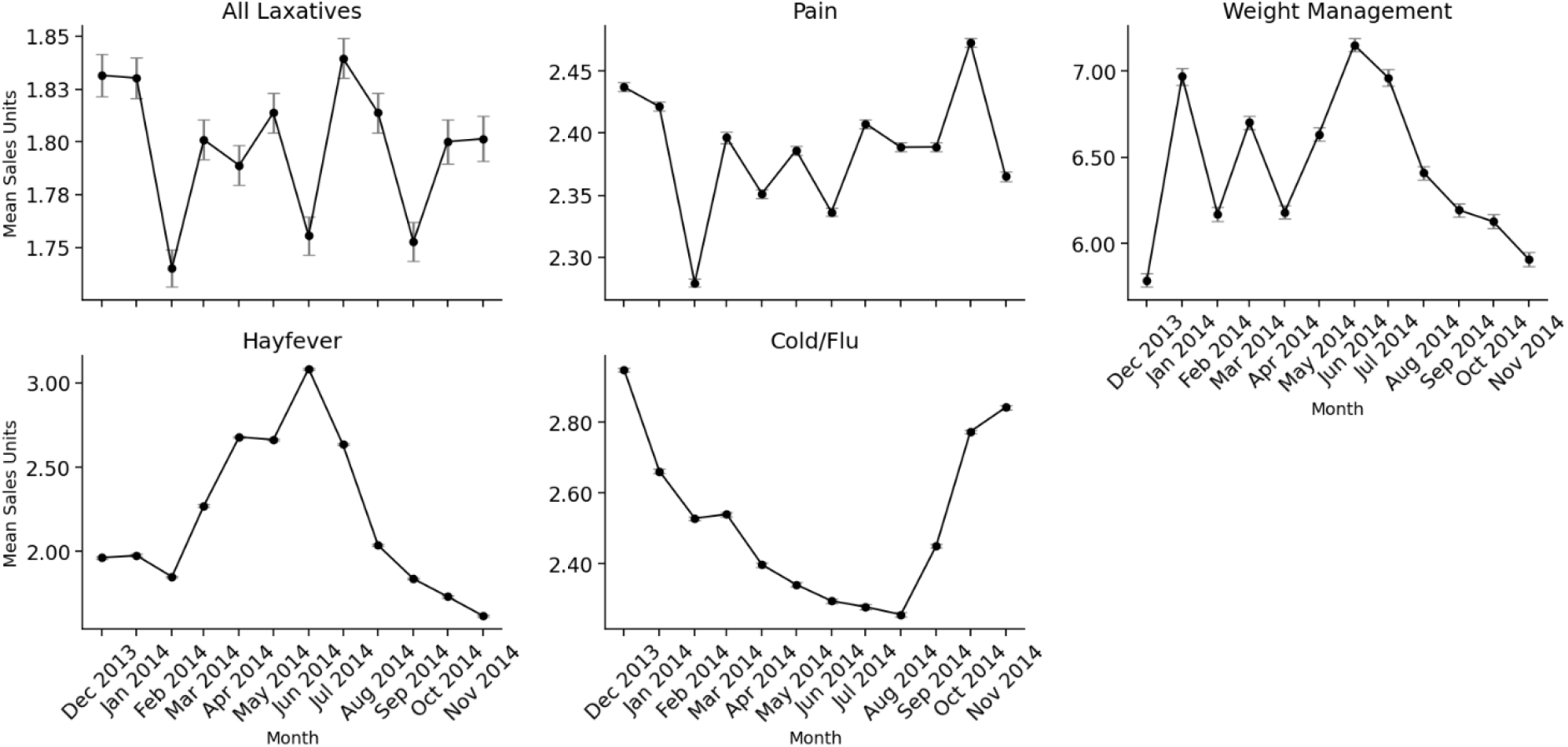
Average monthly sales units (product sales) by product category from December 2013 to November 2014; intervals demonstrate standard errors.

The trends in Figure 1 are as expected. Specifically, sales of hay fever products peak in spring in line with seasonal allergies, cold and flu medication sales peak in the winter months in line with seasonal illnesses, pain product sales remain relatively consistent (with some increase during winter), and weight management product sales increase in the summer months and in the New Year (with a dip in February). For laxatives, the trends are less clear, with fluctuations year-round; there appear to be drops in sales around the start of the year, as well as in June and September.

As a comparison between sales units and dosage, we also plot monthly doses purchased for different laxative products (see Figure 2). This was calculated for the top 10% of buyers of all laxatives (n = 85,578 buyers), stimulants (n = 60,685), and non-stimulants (n = 20,109) separately.

**Fig 2:**
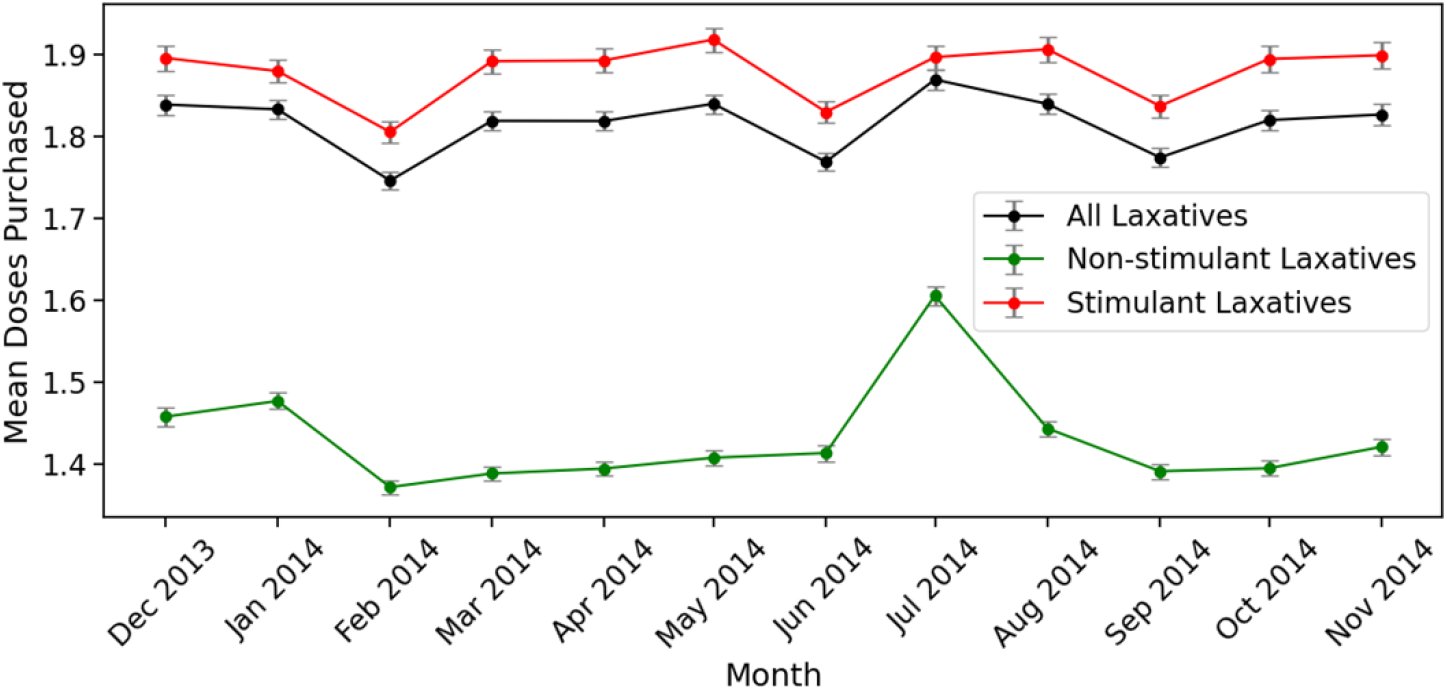
Average monthly laxative doses by laxative type, purchased from December 2013 to November 2014; intervals demonstrate standard errors.

For stimulant-based laxatives, we observe a slow increase in purchasing through the summer, decreasing again in September, and increasing in the lead up to Christmas; we also see a decrease in purchasing at the start of the year. For non-stimulants, we see a large spike in July, as well as an increase around the New Year. The trends for overall laxative purchasing largely mirror those for stimulants, reflecting the high proportion of sales compared to non-stimulants.

### 2.2. Regression Analyses

#### 2.2.1. Seasonal Variation in Stimulant Laxative Purchases

We tested whether there was seasonal variation in stimulant laxative purchases, comparing doses purchased in January and December (H1a), as well as in summer months and September (H1b). Results are shown in Table 3.

There is no evidence for a difference in stimulant laxative purchasing between December and January (H1a). However, there is evidence that more stimulant doses were purchased in both July and August compared to September, with no effect in June, and fewer doses purchased in May compared to September (H1b). This partially supports our hypothesis. In both models, the significance of the inflation constant and alpha justify the use of the model.

#### 2.2.2. Seasonal Variation in Laxative Purchases, Modulated by Laxative Type

We tested whether seasonal variation in laxative purchasing was dependent on the laxative type (stimulant versus non-stimulant), comparing doses purchased in January and December (H2a), as well as in summer months and September (H2b). Results are shown in Table 4.

In both models, we found evidence that more stimulant doses were purchased overall compared to non-stimulants. There is evidence that more doses were purchased in January compared to December, and—contrary to our hypothesis—this trend is stronger for non-stimulants (H2a). There is also evidence that more doses were purchased in May and August compared to September, and this trend is again stronger for non-stimulants (especially in May) (H2b). No seasonal effects were found for June and July. In both cases, the significance of the inflation constant and alpha justify the use of the model.

#### 2.2.3. Seasonal Variation in Weight Management Product Purchases

Finally, we tested whether there was seasonal variation in general diet and weight management products, comparing trends between January and December (H3a), as well as between the summer months and September (H3b). This was tested for the top 10% of buyers of weight loss products (n = 109,827), covering 3,338,922 purchases. Of these, 2990 customers also bought a laxative product (of any type). Among this subgroup, there was a weak positive Pearson correlation (r = 0.11) between the total number of weight-related and laxative products purchased, suggesting a modest tendency for co-use among individuals who purchased both.

Regression results are shown in Table 5.

These results show that more weight management products were purchased in January compared to December, and more were purchased in all summer months compared to September, supporting both hypotheses (H3a, H3b).

## 3. Discussion

This study examined seasonal patterns in laxative purchasing, focusing on whether trends align with seasonal motivations for weight loss and sales of weight management products. Using large-scale, real-world transaction data from a major UK pharmacy retailer (Dec 2013 to Dec 2014), we explored how laxative use varied across the New Year and summer periods. Laxatives were categorised into stimulant or non-stimulant products based on their active ingredients, and *maximum dose duration* was calculated for individual products to estimate the number of doses purchased by an individual. Analyses focused on the top 10% of buyers to identify habitual use.

We hypothesised that stimulant laxative purchases would increase in January compared to December (H1a), reflecting New Year body image concerns, and that this trend would be stronger for stimulants compared to non-stimulants (H2a), due to their perceived weight loss effects. Although we found no difference between January and the December prior with a simple comparison, sales were higher in January when controlling for laxative type. Contrary to the H2a, this increase was more pronounced for non-stimulant laxatives, suggesting that stimulant use was less sensitive to monthly change. We also hypothesised increased stimulant purchases during summer vs. September (H1b), with a stronger effect for stimulants (H2b). More stimulant doses were purchased in July and August compared to September, although fewer were purchased in May. After controlling for laxative type, the August effect persisted, but the July effect disappeared and the May effect reversed. Again, this trend was stronger for non-stimulants, contrary to our hypothesis. Together, these results reflect seasonal increases in laxative purchases during January (compared to December) and August (compared to September), which may reflect motivations linked to body image. During the summer, social pressures and seasonal activities associated with warmer weather heighten body dissatisfaction and weight-loss motivation [20], while the New Year brings similar societal expectations to lose weight [21].

We also predicted similar seasonal trends in weight management products. Purchases increased in January compared to December (H3a), and were higher in summer compared to September (with strongest effects in June and July). This is consistent with seasonal weight loss reported in the literature [13], and likely reflects the increased focus on body image as temperatures rise [20]. These patterns mirror those seen in laxative sales, which may suggest that both categories are influenced by weight loss concerns, although product purpose—weight loss versus digestive health—may account for some differences. In contrast, control categories (hay fever, pain, cold medications) followed expected seasonal trends, emphasising the unique fluctuations in weight management and laxative products.

### 3.1. Dosage vs. Sales Units

We used *maximum dosage duration* as the outcome variable to account for differences in pack size across laxative products (e.g., 12 vs. 48 tablets), offering a more accurate measure of use than units sold. This is particularly important when investigating usage trends, as it reveals whether consumers are using higher or lower doses of a product regardless of how many units they purchase.

By focusing on dosage rather than sales, we were able to identify seasonal variations that might not be apparent when looking only at sales. Regression results using units sold (available in the Supplementary Material) were largely consistent, supporting the same hypotheses (H1a–H2b). Both outcomes showed increased stimulant laxative purchases in January vs. December (H1a, H2a), with stronger effects for stimulants. Similarly, stimulant use was higher in summer than September (H1b), though specific month effects varied (e.g., a July effect appeared in units sold but not dosage). Overall, dosage-based measures provided a more accurate measure of purchasing behaviour, potentially offering more precise insights into self-medication trends compared to sales units.

Unfortunately, implementing this approach across all product categories in this study was not feasible due to the large number of individual items: 121 hay fever products, 447 weight management items, 449 pain relief products, and 409 cold/flu medications. However, future research could benefit from incorporating dosage data to provide more accurate insights into self-medication (e.g., leveraging natural language processing techniques to facilitate this on a larger scale).

### 3.2. The Use of Loyalty Card Data to Inform Policy

Recent studies have highlighted the potential of loyalty card transactions for evaluating public health policy. For example, prior studies have assessed the impact of the sugar tax in the UK and Spain [22, 23], the effectiveness of a salt reduction campaign in the UK [24], and the impact of alcohol policy reform in Finland [25]. However, no studies have yet used loyalty card data to evaluate policies related to self-medication.

In 2020, the Medicines & Healthcare products Regulatory Agency (MHRA) in the UK introduced new regulations to curb stimulant laxative abuse [26]—reducing pack sizes, adding on-pack warnings about weight loss use, and restricting over-the-counter sales to individuals over 18. However, until now, there has been no data-driven evidence to support the implementation of these measures; the evidence base at the time largely relied on case reports and clinical insight.

Our study retrospectively supports the rationale for regulation. We identified seasonal spikes in stimulant laxative purchases, especially during periods commonly associated with weight loss efforts (e.g., summer). Additionally, 2,990 customers purchased both a weight management and laxative product, with a weak correlation indicating minimal degree of co-purchasing. However, this should be interpreted cautiously, as we cannot confirm actual use or intent behind purchasing. While seasonal factors such as travel-related illness or dehydration could also influence laxative use, we strengthen our interpretation by comparing with unrelated medication categories. Taken together, the data suggest that some laxative use may be plausibly linked to misuse with weight management intentions, providing empirical support for the rationale behind the policy change. It also highlights an opportunity for future studies to evaluate seasonal trends in laxative use post-regulation to evaluate the impact of the policy.

### 3.3. Future Research

Our study addresses an important public health concern, providing insights into real-world laxative purchasing behaviour across a large sample of consumers. By focusing on the top 10% of buyers, we effectively quantify the behaviours of the most habitual laxative users. Further, by translating our outcome to reflect dosage rather than sales units, we allow for a more accurate reflection of consumption. These methodological choices open up new avenues to use digital footprint data to understand unobserved health conditions.

Our work includes some limitations. Firstly, our analyses may reflect misalignment between shopping behaviour and consumption. For example, one person may buy a 60-day supply in May, while another buys two 30-day supplies in May and June—despite identical consumption. This discrepancy could bias interpretations of monthly sales trends; this may be addressed in future work by incorporating time-series models. Moreover, standardising product sales by dosage introduced some assumptions about consumer behaviour. In particular, our approach relied on the *maximum dose duration* for an adult; it is possible that not all consumers adhere to this dosage (e.g., due to age or personal preferences). Finally, future studies should incorporate linkages to cross-sectional surveys or medical records to delineate motivations for laxative use. The presence of demographic information will provide ground truth, allowing researchers to more accurately infer conditions which may have influenced laxative purchasing (e.g., eating disorders, digestive issues).

Future work should also aim to delineate the effect of the festive period on laxative purchasing patterns. More broadly, there is a need to develop methods to account for purchasing spikes driven by seasonal or cultural events (e.g., Christmas), promotional activities (e.g., Black Friday), or otherwise. These reflect temporary trends which create short-term surges in sales, skewing results. Developing techniques to adjust for these factors will help provide a more accurate understanding of consumer behaviour. Further studies are needed to identify the impact of the MHRA regulation on UK stimulant laxative purchasing. In addition, comparative studies across different countries with varying policies (e.g., restrictions, regulations) would be valuable to identify effective strategies to combat laxative misuse and identify best practices.

### 3.4. Conclusion

This study provides novel insights into seasonal trends in laxative purchasing, revealing potential links between stimulant laxative use and weight-related motivations, particularly during the summer months. By leveraging large-scale transaction data and focusing on dosage rather than sales units, we offer an objective and accurate representation of laxative consumption patterns. Our findings suggest that stimulant laxative purchases fluctuate in ways similar to weight management products, reinforcing concerns about their misuse for non-medical reasons. Further research is needed to examine post-regulation purchasing patterns to assess whether the MHRA measures have effectively reduce stimulant laxative misuse. Additionally, integrating transaction data with medical records or demographic surveys could help distinguish between legitimate medical use and weight management motivations, informing targeted interventions to address misuse.

## 4. Materials and Methods

### 4.1. Transaction Data

The dataset used in this study comprises individual customers’ purchasing histories from a major UK health and beauty retailer, linked through their loyalty cards at the time of purchase, spanning December 2013 to December 2014. Data covers 2568 stores across the UK. Each transaction includes the sales units (number of items purchased), quantity, price, time and date of purchase, and identifier codes for both items and stores. The records were provided to researchers fully-anonymised.

The complete retailer dataset includes 2,702,449 customers (n = 234,938,722 purchases), approximately 87% of whom are female. Of these, 748,375 participants purchased a laxative product during the specified timeframe, after excluding anomalous purchases. Anomalous purchases were defined as purchases with negative sales units (n = 43,677), which represent refunds, or those with sales units exceeding 20 (n = 4), an unusually high number likely due to data entry errors.

The study design was approved by University of Nottingham Computer Science Ethics Committee.

### 4.2. Data Pre-processing

The dataset was prepared by: (1) categorising laxative products into stimulants versus non-stimulants, (2) calculating the *maximum dosage duration* contained within each product, and (3) limiting to most frequent laxative buyers. The process for this is described below.

#### 4.2.1. Categorising Stimulants versus Non-Stimulants

Laxative products were categorised into three groups: stimulant (denoted as a 1), non-stimulant (0), or other (2). There were 105 unique laxative products: 50 stimulant, 47 non-stimulant, and 8 categorised as other/undefined.

Products were manually defined according to the primary ingredient in the product. Stimulant medications were defined as those whose the primary ingredients were: sodium picosulfate (e.g., Dulcolax liquid), bisacodyl (e.g., Dulcolax tablets, retailer own-brand tablets), and sennoside (e.g., Senna tablets, Senokot products). Non-stimulants were defined as those where the primary ingredients were: ispaghula husk (e.g., Fybogel), paraffin (e.g., liquid paraffin), lactulose, sterculia (e.g., Normacol), sodium citrate (e.g., Micralax), docusate sodium (e.g., DulcoEase capsules), macrogol (e.g., Movicol sachets), guarana (e.g., supplement), and figs (e.g., syrup of figs supplement).

The other/undefined category included non-medication products (like suppository applicators and Epsom salts), as well as those whose description was not sufficient to identify the product and its primary ingredient; these products were excluded from analyses.

#### 4.2.2. Defining Maximum Dosage Duration

Previous works have used direct sales unit data to track medication purchases over time [9, 10]; however, such metrics, in this instance, may lead to inaccurate inferences given the heterogeneity across medication products in terms of number of doses per product. Specifically, if an individual purchases only one box of laxatives per month, the transaction data would reflect a single purchase regardless of the quantity contained in the box (e.g., a pack of 12 tablets versus a pack of 24). As a result, an actual increase in laxative intake could go unnoticed, as the transaction data does not account for the variation in dosage or quantity per pack.

To address this, we defined a standardised variable *maximum dosage duration* for all laxative products, which describes the total number of days’ worth of medication contained within the product, according to the maximum recommended adult dosage. The measure is defined as:

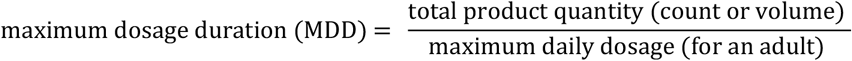

Total product quantity was provided within the item descriptions in the transaction data, while dosage information was collected via manual searches. We used a variety of databases to inform our research, including: the retailer’s own website, product specific websites (e.g., https://www.senokot.co.uk/), and online pharmacies (e.g., chemist-4-u). Table 1 provides some example products with their quantity, maximum daily dosage, and maximum total daily dosage.

**Table 1:**
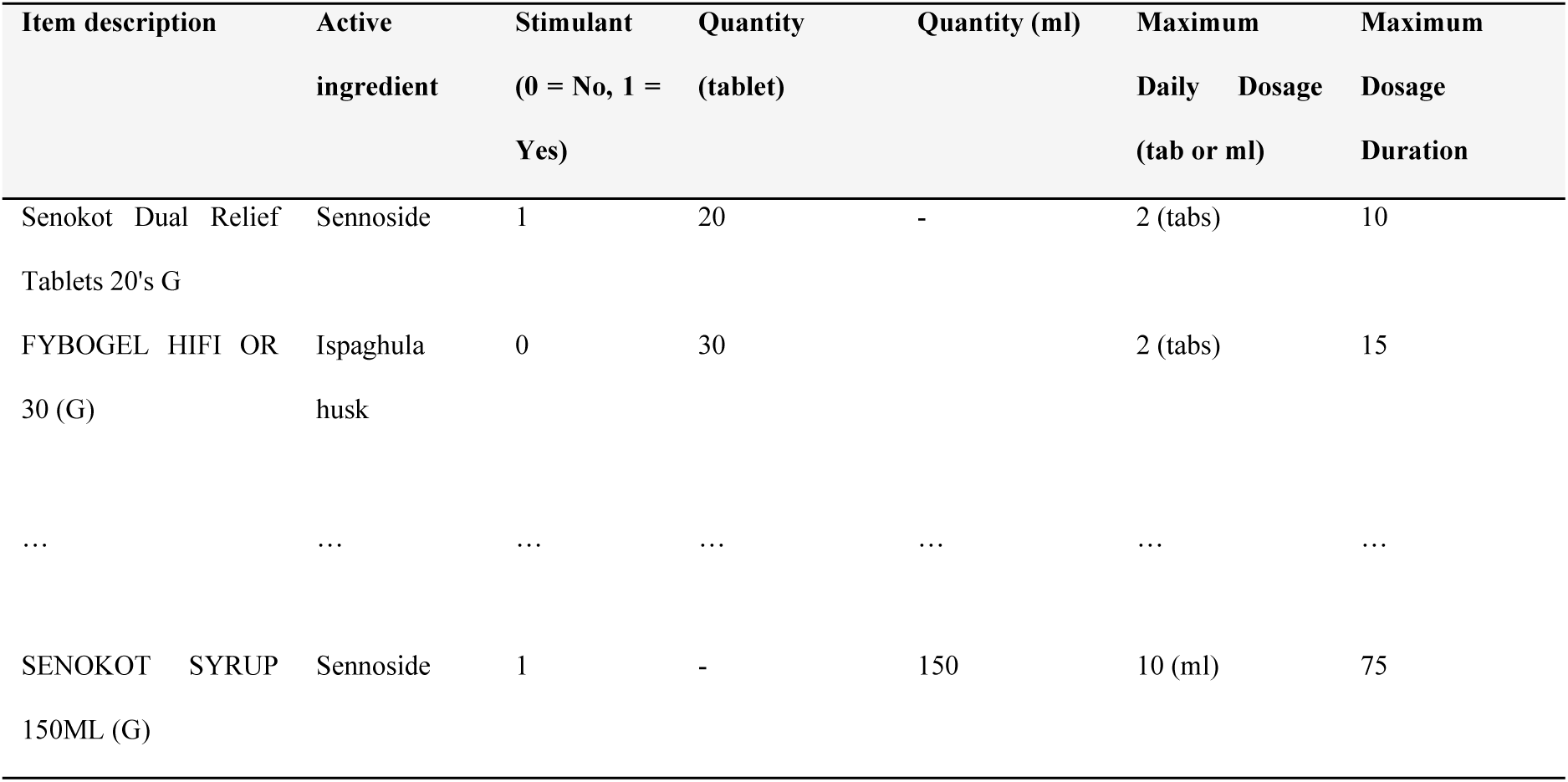
Example product categorisation and dose quantities.

Standardising dosage in this way allows comparison between different forms of the same medication (e.g., between liquid and tablets), and could be extended to enable comparisons between entirely different types of medications (e.g., painkillers against flu medications).

It should be noted that while dosage (rather than sales units) are used in this study to represent laxative sales, this was only possible given the small number of products in the dataset (n = 105 unique products), which allowed for manual labelling of individual products. Ideally, this would have been replicated in our control categories (e.g., hay fever), but was not possible due to the size of those datasets; sale units were instead used in these cases. However, this does not compromise the validity of the comparison, as sales patterns across different medication types are used only to contextualise the laxative data at an aggregate level.

#### 4.2.3. Frequent Buyers as a Proxy for Habitual Users

Previous studies have highlighted the need to identify regular customers when evaluating transaction data, given their ability to demonstrate more consistent purchasing behaviour [14]. This consistency helps reduce noise in analyses by focusing on patterns more likely to reflect genuine consumer preferences rather than one-off or irregular purchases.

Previous approaches include limiting analyses to participants with self-reported loyalty above 41% [15, 16], incorporating loyalty as a control variable [17], or refining datasets to frequent shoppers based on visits or weekly expenditure [18]. Frequent laxative buyers may reflect habitual purchasing for managing ongoing health concerns (e.g., digestive issues); we suggest that high purchasing levels may be due to misuse in relation to weight management, particularly among stimulant laxative users. As such, we limit our analyses to the top 10% of laxative buyers based on maximum dosage duration purchased between December 2013 and December 2014. This threshold was chosen to balance maximising our dataset while focusing on individuals with the highest purchasing behaviour.

In a sensitivity analysis, we also repeated hypotheses H1a-H2b using all buyers (see Supplementary Material).

### 4.3. Statistical Approach

Each hypothesis is tested using a regression analysis, as summarised in Table 2. The outcome for hypotheses H1a– H2b is the *maximum dosage duration* of either stimulant laxatives (H1a, H1b) or all laxatives (H2a, H2b) purchased by an individual within a given month; for simplicity, this is referred to as “doses purchased”. For hypotheses H3a–H3b, the outcome is the number of sales units for diet and weight management products.

**Table 2:**
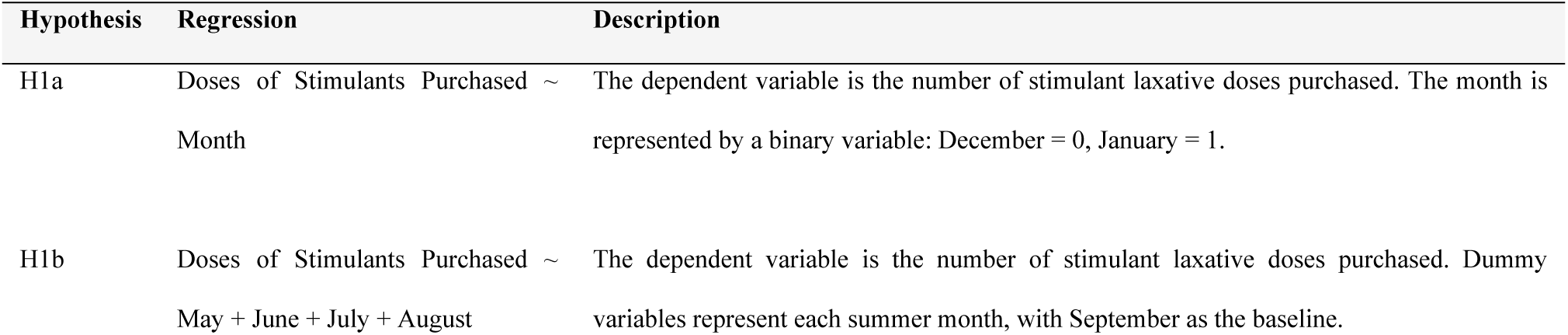
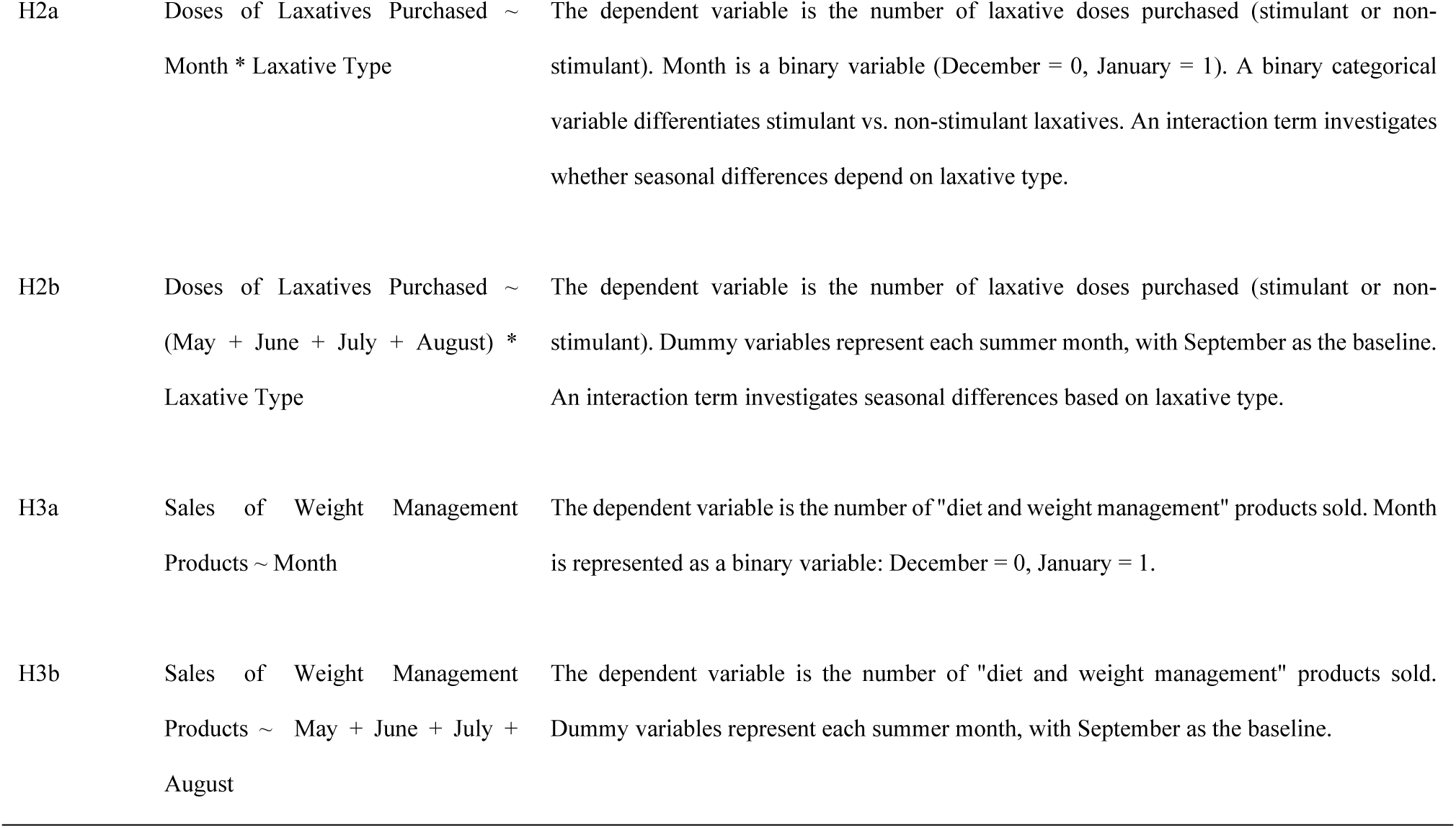
Hypotheses with regression models and explanations.

| Hypothesis | Regression | Description |
| --- | --- | --- |
| H1a | Doses of Stimulants Purchased ~ Month | The dependent variable is the number of stimulant laxative doses purchased. The month is represented by a binary variable: December = 0, January = 1. |
| H1b | Doses of Stimulants Purchased ~ May + June + July + August | The dependent variable is the number of stimulant laxative doses purchased. Dummy variables represent each summer month, with September as the baseline. |
| H2a | Doses of Laxatives Purchased ~ Month * Laxative Type | The dependent variable is the number of laxative doses purchased (stimulant or non-stimulant). Month is a binary variable (December = 0, January = 1). A binary categorical variable differentiates stimulant vs. non-stimulant laxatives. An interaction term investigates whether seasonal differences depend on laxative type. |
| H2b | Doses of Laxatives Purchased ~ (May + June + July + August) * Laxative Type | The dependent variable is the number of laxative doses purchased (stimulant or non-stimulant). Dummy variables represent each summer month, with September as the baseline. An interaction term investigates seasonal differences based on laxative type. |
| H3a | Sales of Weight Management Products ~ Month | The dependent variable is the number of "diet and weight management" products sold. Month is represented as a binary variable: December = 0, January = 1. |
| H3b | Sales of Weight Management Products ~ May + June + July + August | The dependent variable is the number of "diet and weight management" products sold. Dummy variables represent each summer month, with September as the baseline. |

**Table 3:**
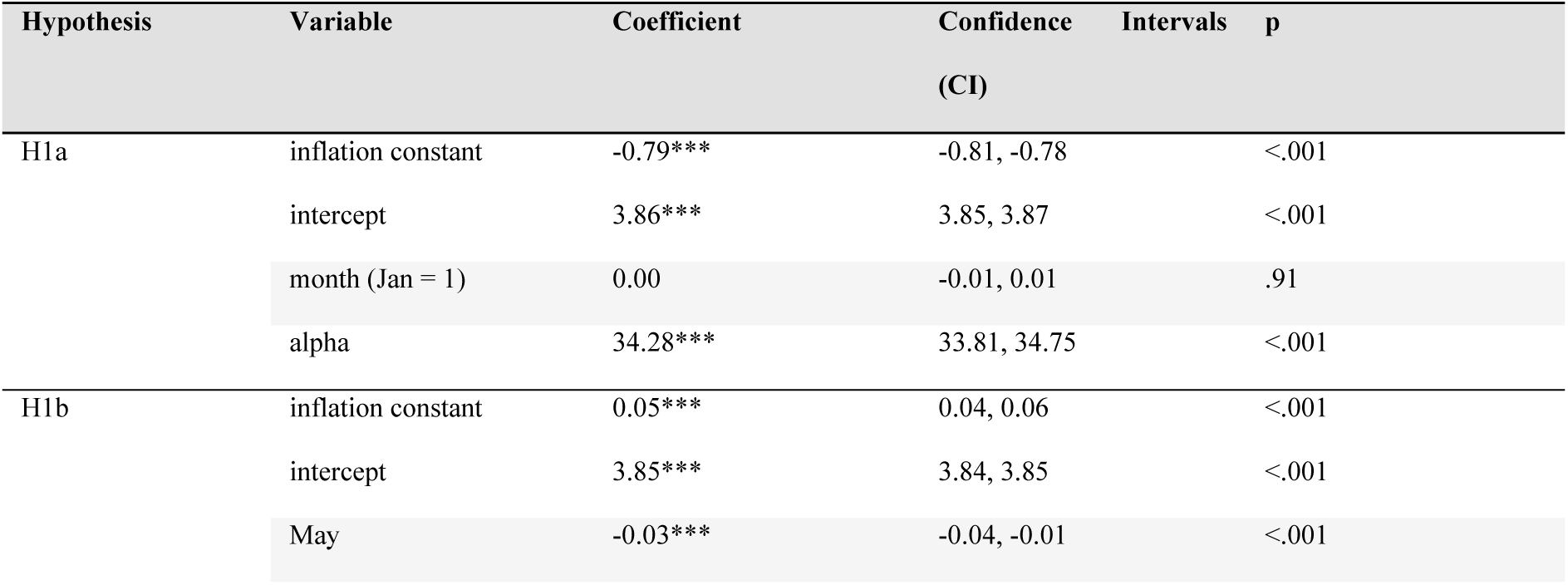
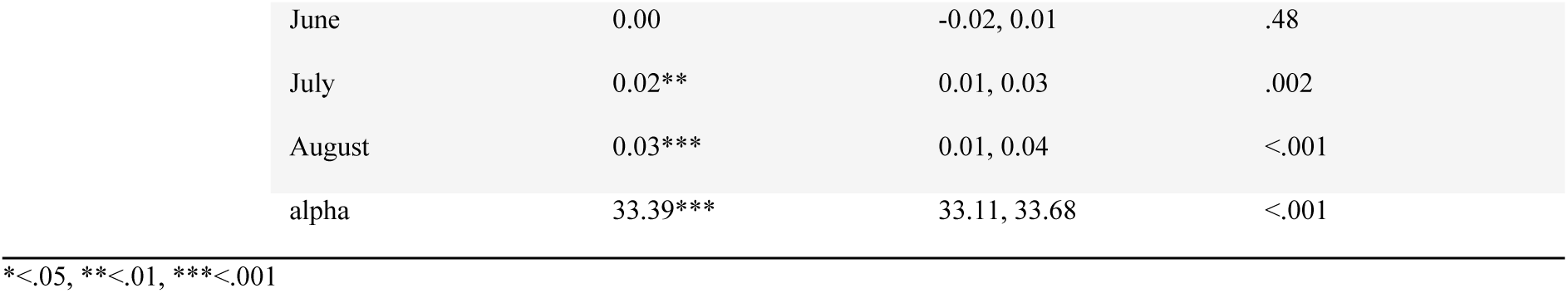
Regression results for H1a and H1b.

| Hypothesis | Variable | Coefficient | Confidence Intervals (CI) | p |
| --- | --- | --- | --- | --- |
| H1a | inflation constant | -0.79*** | -0.81, -0.78 | <.001 |
|  | intercept | 3.86*** | 3.85, 3.87 | <.001 |
|  | month (Jan = 1) | 0.00 | -0.01, 0.01 | .91 |
|  | alpha | 34.28*** | 33.81, 34.75 | <.001 |
| H1b | inflation constant | 0.05*** | 0.04, 0.06 | <.001 |
|  | intercept | 3.85*** | 3.84, 3.85 | <.001 |
|  | May | -0.03*** | -0.04, -0.01 | <.001 |

|  |  |  |  |
| --- | --- | --- | --- |
| June | 0.00 | -0.02, 0.01 | .48 |
| July | 0.02** | 0.01, 0.03 | .002 |
| August | 0.03*** | 0.01, 0.04 | <.001 |
| alpha | 33.39*** | 33.11, 33.68 | <.001 |

\*<.05, \*\*<.01, \*\*\*<.001

**Table 4:**
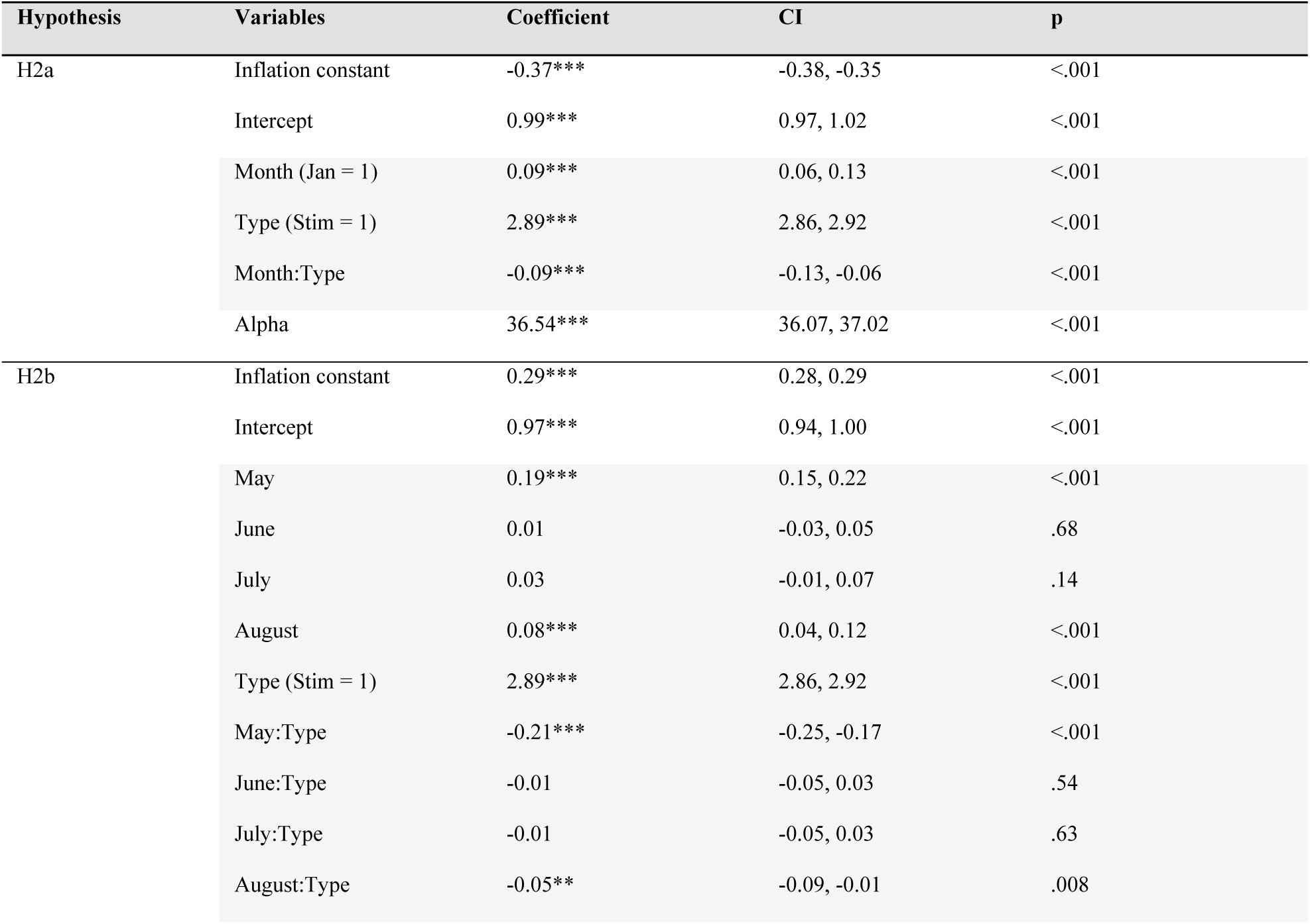
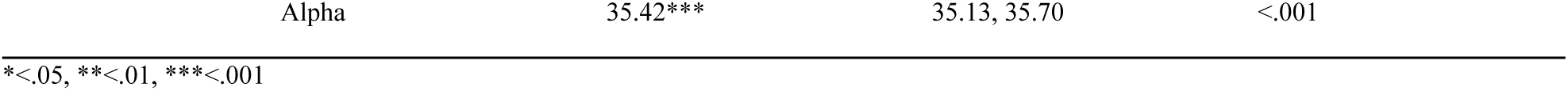
Regression results for H2a and H2b.

**Table 5:** Regression results for H3a and H3b.

|  | Variable | Coefficient | CI | P |
| --- | --- | --- | --- | --- |
| H3a | Inflation constant | -3.89*** | -4.37, -3.40 | <.001 |
|  | Intercept | 1.33*** | 1.31, 1.34 | <.001 |
|  | Month (Jan = 1) | 0.34*** | 0.32, 0.35 | <.001 |
|  | Alpha | 7.90*** | 7.75, 8.04 | <.001 |
| H3b | Inflation constant | -0.79*** | -0.81, -0.78 | <.001 |
|  | Intercept | 1.44*** | 1.43, 1.45 | <.001 |
|  | May | 0.15*** | 0.13, 0.16 | <.001 |
|  | June | 0.28*** | 0.27, 0.29 | <.001 |
|  | July | 0.26*** | 0.25, 0.27 | <.001 |
|  | August | 0.09*** | 0.08, 0.11 | <.001 |
|  | Alpha | 7.97*** | 7.89, 8.05 | <.001 |

\*<.05, \*\*<.01, \*\*\*<.001

We use zero-inflated negative binomial regression in this analysis, a model well-suited for count data characterised by overdispersion and an excess of zeros. The negative binomial component accounts for when the variance of the outcome variable significantly exceeds its mean, while the zero-inflation component accounts for the disproportionately high frequency of zero outcomes (e.g., months when customers made no laxative purchases). This approach was chosen because the outcome variable in each regression showed both overdispersion and a substantial proportion of zeros. Model outputs were interpreted using 95% confidence intervals and p-values to evaluate any meaningful effects.

As a comparison, we also repeated hypotheses H1a-H2b using sales units as the outcome (see Supplementary Material).

#### 4.3.1. Pre-Registration

Our hypotheses have been pre-registered [19]. We have made some changes to the methodologies which are noted here:

- We use data from 2013-2014, rather than 2014-2015; this is because, upon inspection, data from 2015 was incomplete. To ensure the use of the most recent and complete dataset, we revised the study period.
- We convert product data into dosage, as opposed to sales, to more accurately capture use, as dosage provides a standardised measure that better reflects potential consumption across products of varying sizes.
- We refine analyses to frequent shoppers only (top 10%). This decision improves the reliability of behavioural inferences, as frequent shoppers offer denser, more consistent data. In contrast, infrequent shoppers may introduce noise due to irregular purchasing or external factors (e.g., shopping elsewhere).

We do not believe these changes to have significantly impacted our conclusions.

## Data Availability Statement

The data underlying this article were provided by a third party under licence / by permission, and so cannot be made available.

## Supplementary Material

### Results for all Laxative Buyers

We tested whether there was seasonal variation in stimulant laxative purchases for <u>all laxative buyers</u> (as opposed to frequent buyers only), comparing doses purchased in January and December (H1a), as well as in summer months and September (H1b). The total number of laxative buyers was 748,375 individuals, comprising 1,683,712 items. The results for both regressions are shown in Table A1.

**Table A1:**
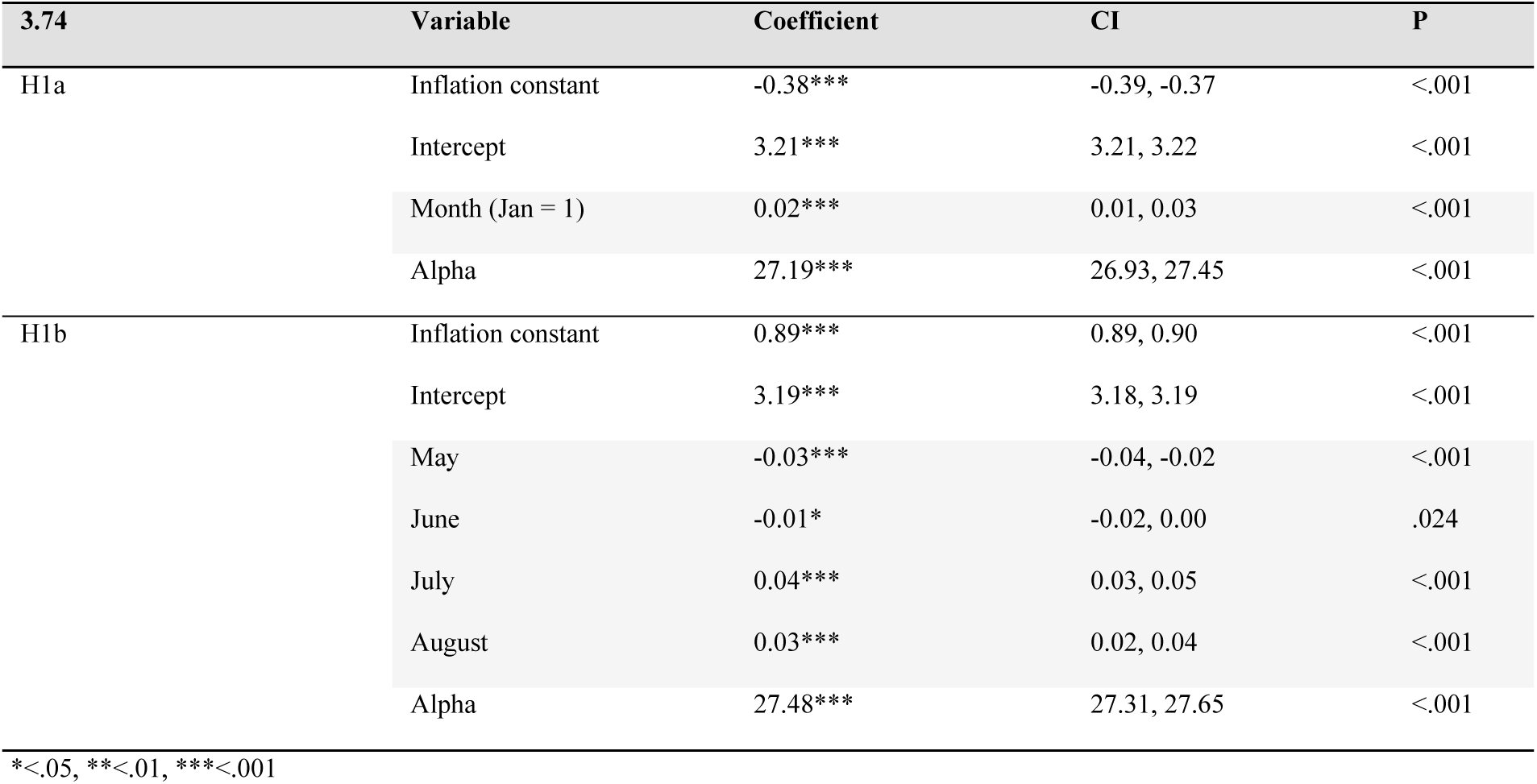
Regression results for H1a and H1b for all laxative buyers.

| 3.74 | Variable | Coefficient | CI | P |
| --- | --- | --- | --- | --- |
| H1a | Inflation constant | -0.38*** | -0.39, -0.37 | <.001 |
|  | Intercept | 3.21*** | 3.21, 3.22 | <.001 |
|  | Month (Jan = 1) | 0.02*** | 0.01, 0.03 | <.001 |
|  | Alpha | 27.19*** | 26.93, 27.45 | <.001 |
| H1b | Inflation constant | 0.89*** | 0.89, 0.90 | <.001 |
|  | Intercept | 3.19*** | 3.18, 3.19 | <.001 |
|  | May | -0.03*** | -0.04, -0.02 | <.001 |
|  | June | -0.01* | -0.02, 0.00 | .024 |
|  | July | 0.04*** | 0.03, 0.05 | <.001 |
|  | August | 0.03*** | 0.02, 0.04 | <.001 |
|  | Alpha | 27.48*** | 27.31, 27.65 | <.001 |

\*<.05, \*\*<.01, \*\*\*<.001

There is evidence that more stimulants were purchased in January compared to December (H1a), matching our findings for frequent buyers. We also found increased purchases in July and August (but reduced in May and June) compared to September (H1b), partially matching our findings for frequent buyers.

**Table A2:**
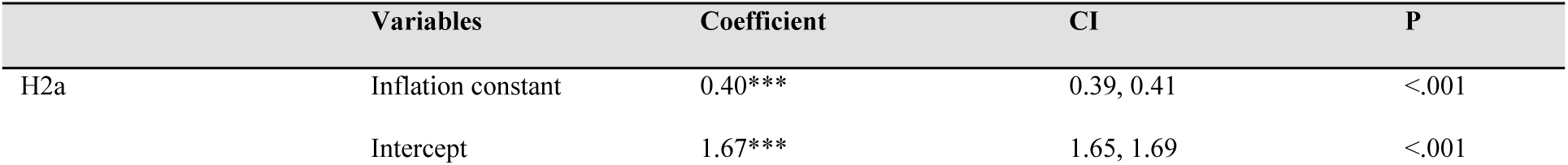
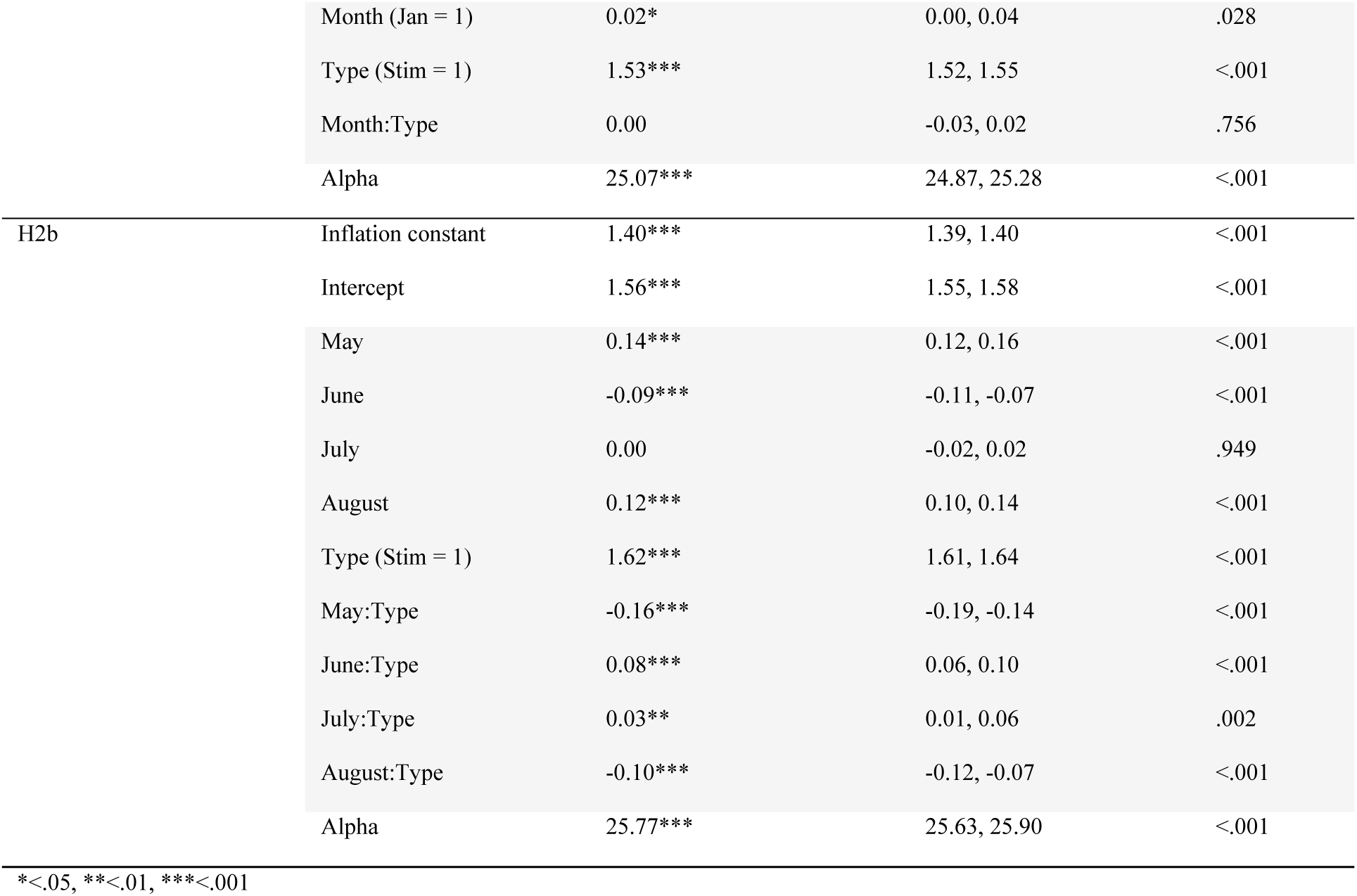
Regression results for H2a and H2b for all laxative buyers.

We also tested whether seasonal variation in laxative purchasing was dependent on laxative type for all laxative buyers, comparing doses purchased in January and December (H2a), as well as in summer months and September (H2b). The results are shown in Table A2.

There is evidence for increased doses purchased in January compared to December, although laxative type shows no effect (H1a)—contrary to our findings for frequent buyers. There is also evidence that more doses were purchased in May and August (but fewer in June) compared to September, and these effects were stronger for non-stimulants for these months (but stronger for stimulants for June and July) (H1b), partially matching our findings for frequent buyers.

### Results using Sales Units

As a point of comparison, we present our regression analyses using sales units as the outcome (as opposed to dosage), again limited to the top 10% of laxative buyers (based on sales units). This gives 85,578 buyers, covering 817,061 items purchased.

We tested whether there was seasonal variation in sales of stimulant laxatives (as opposed to doses purchased), comparing sales in January and December (H1a), as well as in summer months and September (H1b). The results are shown in Table A3.

**Table A3:** Regression results for H1a and H1b using sales units as the outcome.

| 3.74 | Variable | Coefficient | CI | P |
| --- | --- | --- | --- | --- |
| H1a | Intercept | 0.19*** | 0.18, 0.20 | <.001 |
|  | Month (Jan = 1) | 0.02* | 0.00, 0.03 | .042 |
|  | Alpha | 0.51*** | 0.50, 0.51 | <.001 |
| H1b | Inflation constant | -0.01** | -0.02, 0.00 | .003 |
|  | Intercept | 3.70*** | 3.69, 3.71 | <.001 |
|  | May | -0.02* | -0.03, 0.00 | .039 |
|  | June | 0.00 | -0.02, 0.01 | .837 |
|  | July | 0.04*** | 0.02, 0.05 | <.001 |
|  | August | 0.03*** | 0.02, 0.05 | <.001 |
|  | Alpha | 40.21*** | 39.85, 40.57 | <.001 |

\*<.05, \*\*<.01, \*\*\*<.001

We found that sales were increased in January compared to December (H1a), consistent with our findings for dosage (Table 3). We also found that sales in both July and August were higher than in September (H1b) (although sales were lower in May, with no effect in June), matching our findings for dosage.

We also tested whether seasonal variation in laxative purchasing was dependent on the laxative type (stimulant versus non-stimulant), comparing sales in January and December (H2a), as well as in summer months and September (H2b). The results are shown in Table A4.

**Table A4:**
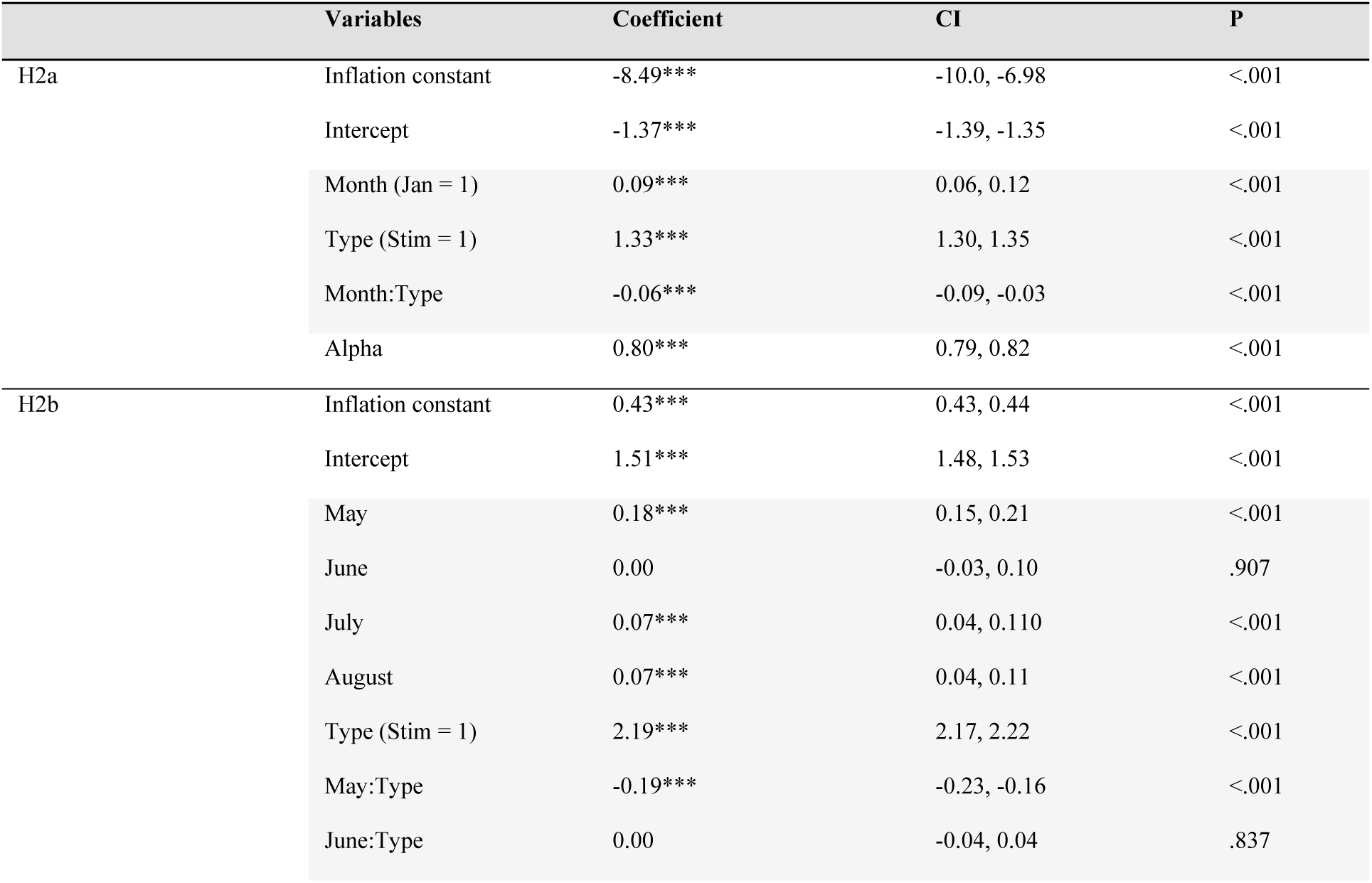
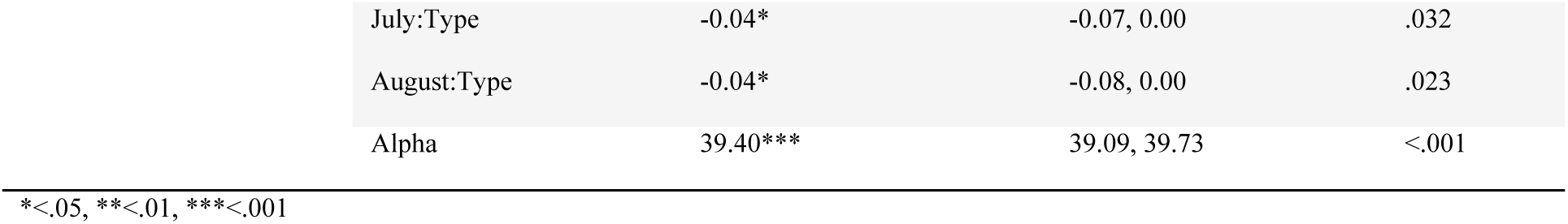
Regression results for H2a and H2b using sales units as the outcome.

|  | Variables | Coefficient | CI | P |
| --- | --- | --- | --- | --- |
| H2a | Inflation constant | -8.49*** | -10.0, -6.98 | <.001 |
|  | Intercept | -1.37*** | -1.39, -1.35 | <.001 |
|  | Month (Jan = 1) | 0.09*** | 0.06, 0.12 | <.001 |
|  | Type (Stim = 1) | 1.33*** | 1.30, 1.35 | <.001 |
|  | Month:Type | -0.06*** | -0.09, -0.03 | <.001 |
|  | Alpha | 0.80*** | 0.79, 0.82 | <.001 |
| H2b | Inflation constant | 0.43*** | 0.43, 0.44 | <.001 |
|  | Intercept | 1.51*** | 1.48, 1.53 | <.001 |
|  | May | 0.18*** | 0.15, 0.21 | <.001 |
|  | June | 0.00 | -0.03, 0.10 | .907 |
|  | July | 0.07*** | 0.04, 0.110 | <.001 |
|  | August | 0.07*** | 0.04, 0.11 | <.001 |
|  | Type (Stim = 1) | 2.19*** | 2.17, 2.22 | <.001 |
|  | May:Type | -0.19*** | -0.23, -0.16 | <.001 |
|  | June:Type | 0.00 | -0.04, 0.04 | .837 |

|  |  |  |  |
| --- | --- | --- | --- |
| July:Type | -0.04* | -0.07, 0.00 | .032 |
| August:Type | -0.04* | -0.08, 0.00 | .023 |
| Alpha | 39.40*** | 39.09, 39.73 | <.001 |

We found evidence that more laxative doses were purchased in January compared to December, and this effect was stronger for non-stimulants, consistent with our findings using dosage (H2a). We also found that more laxatives were purchased in May, July, and August compared to September (H2b), and these effects were weaker in stimulants. This is very similar to our results using dosage, except that there are July effects in this case.

